# SARS-CoV-2 spike T cell responses induced upon vaccination or infection remain robust against Omicron

**DOI:** 10.1101/2021.12.26.21268380

**Authors:** Roanne Keeton, Marius B. Tincho, Amkele Ngomti, Richard Baguma, Ntombi Benede, Akiko Suzuki, Khadija Khan, Sandile Cele, Mallory Bernstein, Farina Karim, Sharon V. Madzorera, Thandeka Moyo-Gwete, Mathilda Mennen, Sango Skelem, Marguerite Adriaanse, Daniel Mutithu, Olukayode Aremu, Cari Stek, Elsa du Bruyn, Mieke A. Van Der Mescht, Zelda de Beer, Talita R. de Villiers, Annie Bodenstein, Gretha van den Berg, Adriano Mendes, Amy Strydom, Marietjie Venter, Alba Grifoni, Daniela Weiskopf, Alessandro Sette, Robert J. Wilkinson, Linda-Gail Bekker, Glenda Gray, Veronica Ueckermann, Theresa Rossouw, Michael T. Boswell, Jinal Bihman, Penny L. Moore, Alex Sigal, Ntobeko A. B. Ntusi, Wendy A. Burgers, Catherine Riou

## Abstract

The SARS-CoV-2 Omicron variant has multiple Spike (S) protein mutations that contribute to escape from the neutralizing antibody responses, and reducing vaccine protection from infection. The extent to which other components of the adaptive response such as T cells may still target Omicron and contribute to protection from severe outcomes is unknown. We assessed the ability of T cells to react with Omicron spike in participants who were vaccinated with Ad26.CoV2.S or BNT162b2, and in unvaccinated convalescent COVID-19 patients (n = 70). We found that 70-80% of the CD4 and CD8 T cell response to spike was maintained across study groups. Moreover, the magnitude of Omicron cross-reactive T cells was similar to that of the Beta and Delta variants, despite Omicron harbouring considerably more mutations. Additionally, in Omicron-infected hospitalized patients (n = 19), there were comparable T cell responses to ancestral spike, nucleocapsid and membrane proteins to those found in patients hospitalized in previous waves dominated by the ancestral, Beta or Delta variants (n = 49). These results demonstrate that despite Omicron’s extensive mutations and reduced susceptibility to neutralizing antibodies, the majority of T cell response, induced by vaccination or natural infection, cross-recognises the variant. Well-preserved T cell immunity to Omicron is likely to contribute to protection from severe COVID-19, supporting early clinical observations from South Africa.

## Main Text

The newest SARS-CoV-2 variant of concern, designated Omicron^1^,was described on 26 November 2021 from sequences from Botswana, Hong Kong and South Africa^2^. Omicron is responsible for the current surge of infections in South Africa, and is becoming globally dominant. With over 30 mutations in the spike protein, a substantial ability to evade the neutralizing antibody response has been described^3-6^. This associates with greater capacity for reinfection^7^, as well as lower early estimates of vaccine effectiveness against symptomatic disease^8,9^. SARS-CoV-2-specific T cells play a key role in modulating COVID-19 severity^10-12^, and provide protective immunity in the context of suboptimal antibody titers^13^. Given Omicron’s extensive ability to escape antibody responses, we determined whether T cells generated in response to vaccination or previous SARS-CoV-2 infection could cross-recognize Omicron.

We examined T cell responses in participants who had received one or two doses of the Ad26.COV2.S vaccine (Johnson and Johnson/Janssen, n = 20/group), two doses of the BNT162b2 mRNA vaccine (Pfizer–BioNTech, n = 15), or who had recovered from infection (n = 15) (**Fig 1a, Supplementary Table 1** and **2**).

**Fig.1:**
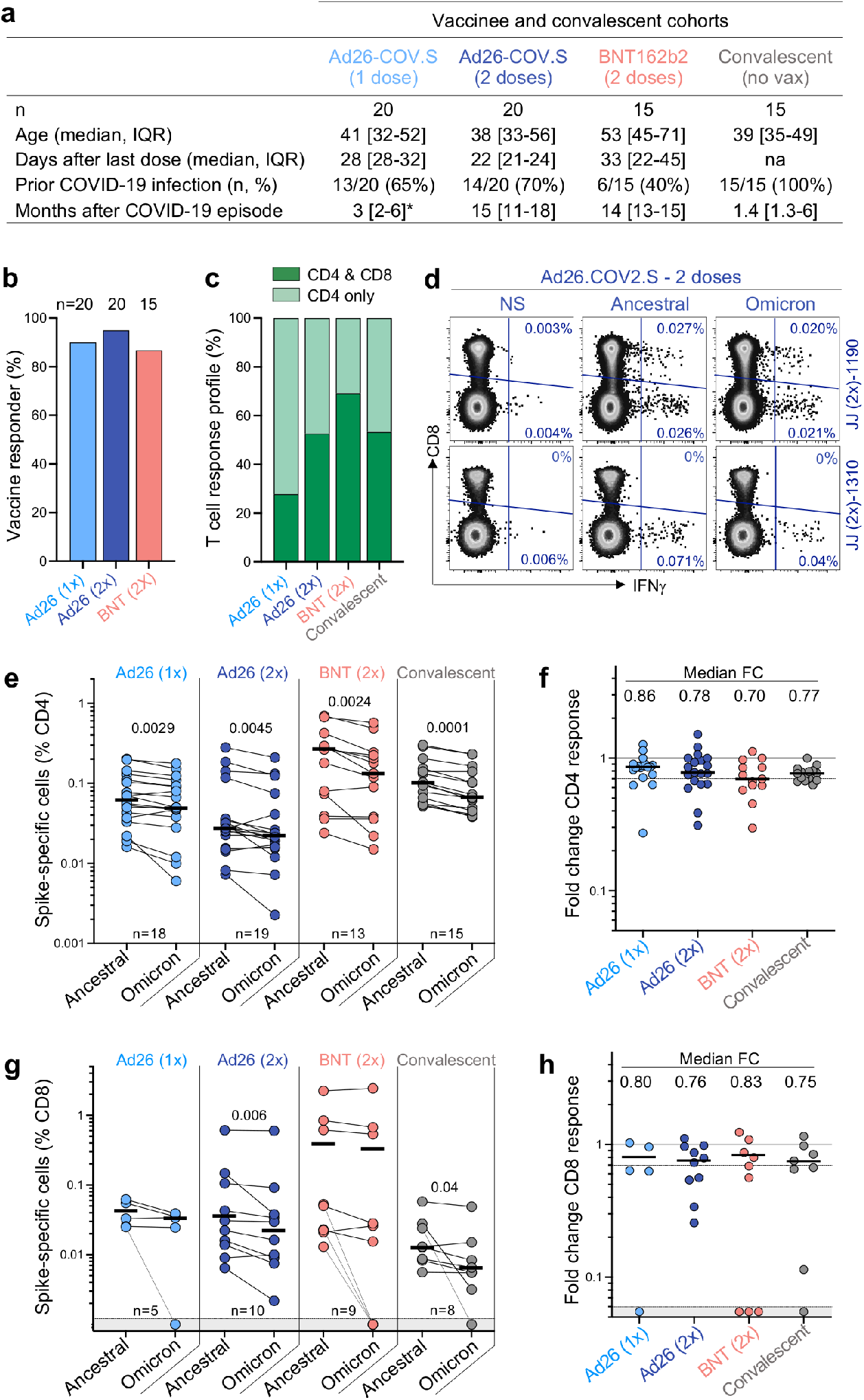
T cell response to the ancestral and Omicron SARS-CoV-2 spike after vaccination and in unvaccinated COVID-19 convalescent patients. **a**, Clinical characteristics of the study groups. *: data regarding time post Covid-19 infection were available for only 6 out of the 13 participants who received one dose of Ad26.COV2.S. **b**, Proportion of participants exhibiting an ancestral spike-specific CD4 T cell response after vaccination with one or two doses of Ad26.COV2.S or two doses of BNT162b2. **c**, Profile of the ancestral spike-specific T cell response in vaccinees and convalescent individuals. **d**, Representative examples of IFN-γ production in response to ancestral and Omicron spike in two individuals who received two doses of Ad26.COV2.S. **e,g**, Frequency of spike-specific CD4 (**e**) and CD8 T cells (**g**) producing any of the measured cytokines (IFN-γ, IL-2 or TNF-α) in response to ancestral and Omicron spike peptide pools. Bars represent median of responders. Differences between SARS-CoV-2 variants were calculated using a Wilcoxon paired t-test. **f, h**, Fold change in the frequency of spike-specific CD4 (**f**) and CD8 T cells (**h**) between ancestral and Omicron spike responses. Bars represent medians. No significant differences were observed between groups using a Kruskal-Wallis test with Dunn’s multiple comparisons post-test. The number of participants included in each analysis is indicated on the graphs.

Convalescent donors were examined a median of 1.4 months (interquartile range [IQR]: 1.3-6 months) after mild or asymptomatic infection. More than 85% of vaccinees generated a T cell response to vaccination, measured 22-32 days after the last dose (**Fig. 1b**). Both vaccination and infection induced spike-specific CD4 T cell responses, while a CD8 response was less consistently detected (**Fig. 1c**). We measured cytokine production (IFN-γ, IL-2 and TNF-α) by intracellular cytokine staining in response to peptide pools covering the full Wuhan-1 spike protein (ancestral) and the Omicron spike (**Fig. 1d** and **Extended Data Fig 1a**).

CD4 T cell frequencies to Omicron spike were consistently and significantly lower than ancestral spike in all groups tested (**Fig. 1e**). This translated to a median decrease of 14-30% of the CD4 response to Omicron, as demonstrated by fold-change (**Fig. 1f**). Similar results were observed for the CD8 T cell response (**Fig. 1g-h**), where vaccinees who had received two doses of Ad26.COV2.S and convalescent donors demonstrated a significant decrease in the magnitude of Omicron spike-specific CD8 T cells, although the other groups did not. There was a median reduction of 17-25% of the CD8 response to Omicron compared to the ancestral virus. Of note, a fraction of responders (5/32; 15%) exhibited a loss of CD8 T cell recognition of Omicron (**Figure 1g** and **Extended Data Fig. 1b**), likely reflecting specific HLA molecules being adversely affected by mutations in particular CD8 epitopes^14^.

Mutations in variant epitopes have the potential to decrease T cell affinity, which may affect the functional capacity of cells^15^. Thus, we compared the polyfunctional profiles of T cells in vaccinees and convalescent individuals and demonstrate similar capacities for cytokine co-expression across all groups for both ancestral and Omicron-specific T cells (**Extended Data Fig. 2a-b** and **3a-b)**. Notably, there were also no differences in the polyfunctional profiles between ancestral and Omicron spike for either CD4 or CD8 T cells (**Extended Data Fig. 2c** and **3c**), indicating the absence of a functional deficit in cross-reactive Omicron T cell responses. We also compared Omicron spike responses to other variants of concern in Ad26.CoV2.S vaccinees, by testing spike peptide pools corresponding to the viral sequences of the Beta and Delta strains (**Extended Data Fig. 4a**). There were no significant differences in cross-reactive CD4 and CD8 T cell responses between Beta, Delta and Omicron (**Extended Data Fig. 4b**), with the exception of a greater decrease in the Omicron CD4 response compared to Beta in recipients of two doses of Ad26.COV2.S. Of note, while prior SARS-CoV-2 infection in vaccinees associated with a higher frequency of spike-specific T cells (**Extended Data Fig. 5a**), it had no impact on Omicron cross-reactivity (**Extended Data Fig. 5b**). Overall, these results show that CD4 and CD8 T cell recognition of Omicron spike is largely preserved compared to the ancestral strain, and is similar to other variants of concern carrying three times fewer mutations.

The SARS-CoV-2 epidemic in South Africa has been characterized by four virologically distinct infection waves (**Fig. 2b**). This enabled us to compare T cell responses in patients from the current fourth epidemic wave, dominated by Omicron, with those infected in prior waves dominated by ancestral (Wave 1, n =17), Beta (Wave 2, n =16) and Delta (Wave 3, n =16) variants (**Fig. 2a**). In addition to extensive mutations in spike, Omicron has 20 additional mutations in other proteins which could also result in T cell escape. Therefore, we measured the frequency of CD4 and CD8 T cells to ancestral spike (S), nucleocapsid (N) and membrane (M) proteins, all major targets of the T cell response^16^. We studied SARS-CoV-2-infected patients who were hospitalized with COVID-19 (**Fig. 2a**). These recently hospitalized patients, recruited between December 1^st^ and 15^th^, 2021 (n = 19), had no history of prior COVID-19 and were unvaccinated. Omicron infection was confirmed by S-gene target failure (SGTF; 7 patients), or whole genome sequencing (5 patients, underway). Seven swabs were unavailable at the time of hospitalization, however with Omicron accounting for 100% of sequences from South Africa at the time of recruitment (**Fig. 2b**), there was a high probability of Omicron infection.

**Fig. 2:**
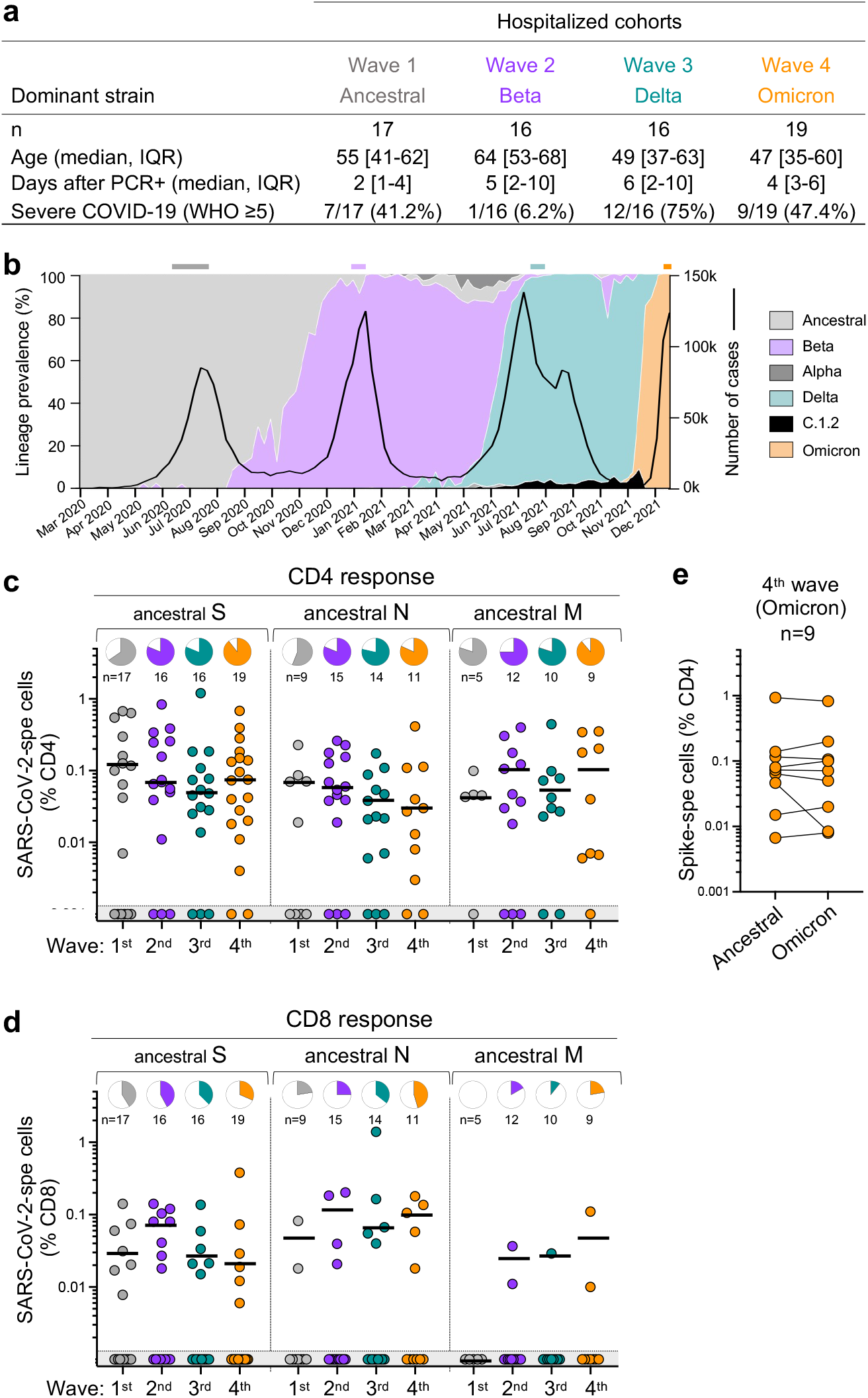
T cell response to ancestral SARS-CoV-2 in unvaccinated hospitalized COVID-19 patients infected with the ancestral, Beta, Delta or Omicron SARS-CoV-2 variants. **a**, Clinical characteristics of the study groups. Severe disease was defined based on oxygen therapy requirement according to the WHO ordinal scale scoring system (O_2_ via high flow to ECMO). **b**, SARS-CoV-2 epidemiological dynamics in South Africa showing the prevalence of different SARS-CoV-2 strains (based on 24,762 sequences; left axis) and the number of COVID-19 cases (right axis). The bars on top of the graph indicate the periods when samples were collected for each epidemic wave. **c,d**, Frequency of SARS-CoV-2-specific CD4 (**c**) and CD8 T cells (**d**) producing any of the measured cytokines (IFN-γ, IL-2 or TNF-α) in response to ancestral SARS-CoV-2 spike (S), nucleocapsid (N) and membrane (M) peptide pools. Pies depict the proportion of participants exhibiting a detectable T cell response to each protein. **e**, Comparison of T cell response to ancestral or Omicron spike in Omicron-infected patients. Bars represent medians of responders. No significant differences were observed between antigens amongst responders using a Kruskal-Wallis test with Dunn’s multiple comparisons post-test. The number of participants included in each analysis is indicated on the graphs.

Despite differences in age, disease severity and co-morbidities across the infection waves (**Fig. 2a** and **Supplementary Table 3**), T cell responses directed at S, N and M in wave 4 patients were of similar magnitude as those in patients infected with other SARS-CoV-2 variants in previous waves (**Fig. 2c** and **d**). The frequency of responders also did not differ markedly across the waves. Furthermore, the magnitude of Omicron spike-specific CD4 responses mounted by Wave 4 patients was highly comparable to ancestral spike (**Fig. 2e**), suggesting that most patients target conserved epitopes in spike.

Overall, these results demonstrate that vaccination and infection induce a robust CD4 and CD8 T cell response that largely cross-reacts with Omicron, consistent with recent work from our laboratory and others on limited T cell escape by Beta, Delta and other variants^17-19^. Despite extensive neutralization escape against Omicron^5^, 70-80% of the T cell response is preserved. The limited effect of Omicron’s mutations on the T cell response suggests that vaccination or prior infection may still provide substantial protection from severe disease. Indeed, South Africa has reported a lower risk of hospitalisation and severe disease compared to the previous Delta wave^20^. Cross-reactive T cell responses acquired through vaccination or infection may contribute to these apparent milder outcomes for Omicron. The resilience of the T cell response demonstrated here also bodes well in the event that more highly mutated variants emerge in the future.

## Data Availability

All data produced in the present study are available upon reasonable request to the authors

---

**Extended Data Fig. 1:**
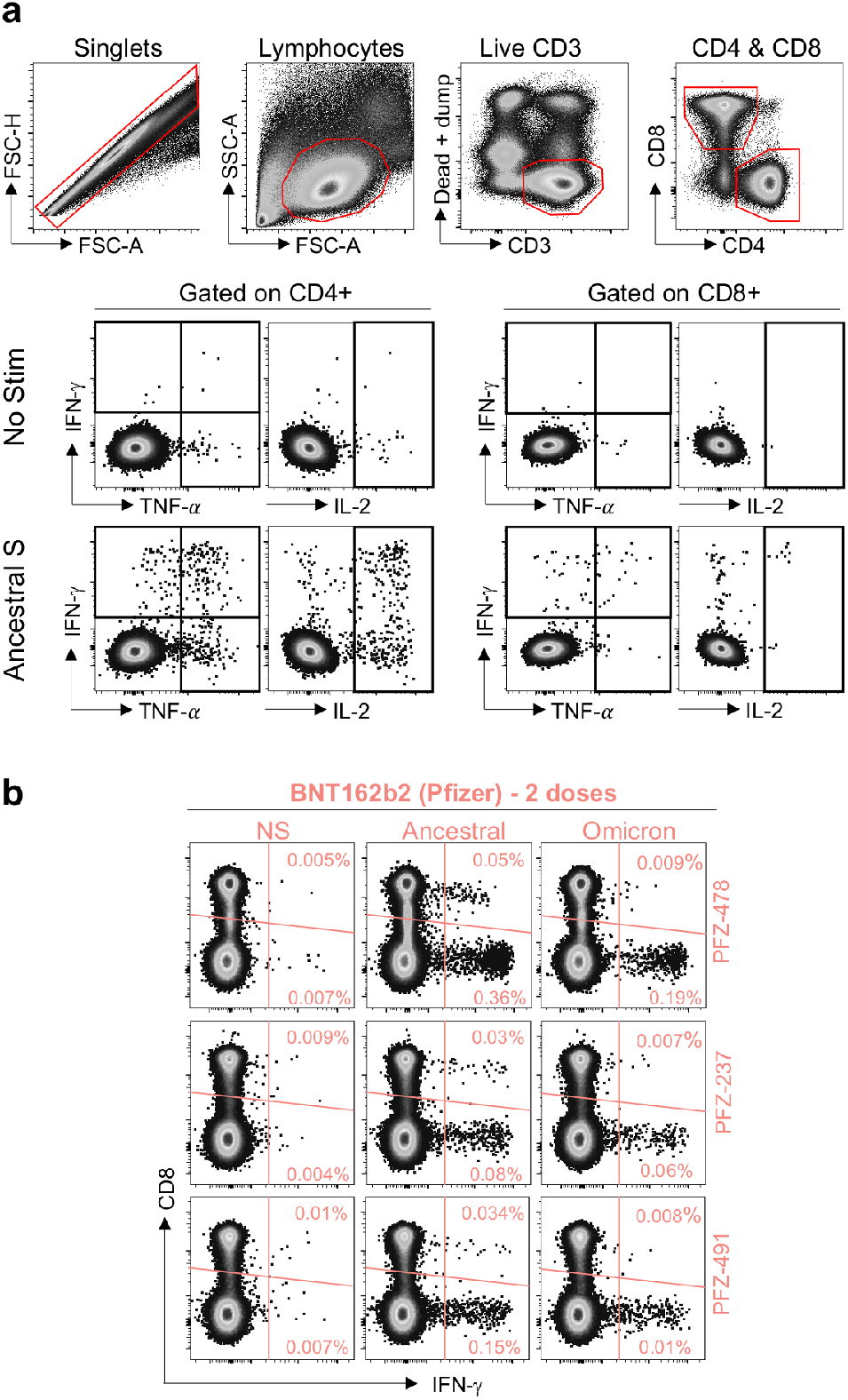
Gating strategy and examples of flow cytometry plots. **a**, Gating strategy and representative examples of SARS-CoV-2 spike-specific IFN-γ, IL-2 and TNF-α production. **b**, Spike-specific expression of IFN-γ in the T cell compartment of the three BNT162b2-vaccinated participants where Omicron-specific CD8 T cells were undetectable.

**Extended Data Fig. 2:**
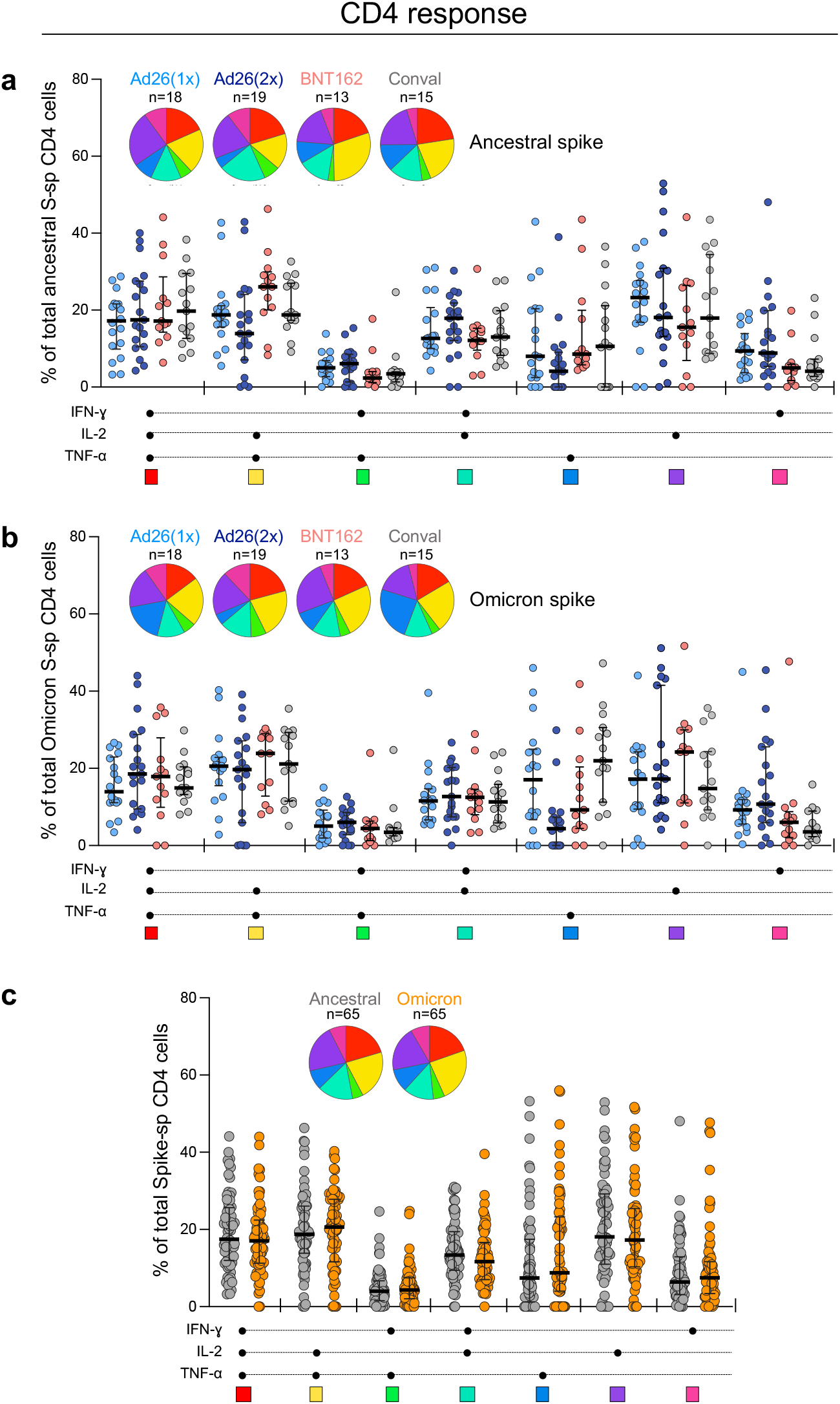
Polyfunctional profiles of SARS-CoV-2-specific CD4 T cells after vaccination and in unvaccinated convalescent volunteers. **a, b**, Comparison of the polyfunctional profile of ancestral (**a**) and Omicron (**b**) spike-specific CD4 T cells between the four groups (Ad26.COV2.S-one dose, Ad26.COV.S-two doses, BNT162b2-two doses and unvaccinated convalescent volunteers). **c**, Comparison of the polyfunctional profile between ancestral and Omicron spike-specific CD4 T cells including all CD4 T cell responding participants, irrespective of their clinical grouping. The medians and IQR are shown. Each response pattern (i.e., any possible combination of IFN-γ, IL-2 or TNF-α expression) is color-coded, and data are summarized in the pie charts. No significant differences were observed between pies using a permutation test. The number of participants included in each analysis is indicated on the graphs.

**Extended Data Fig. 3:**
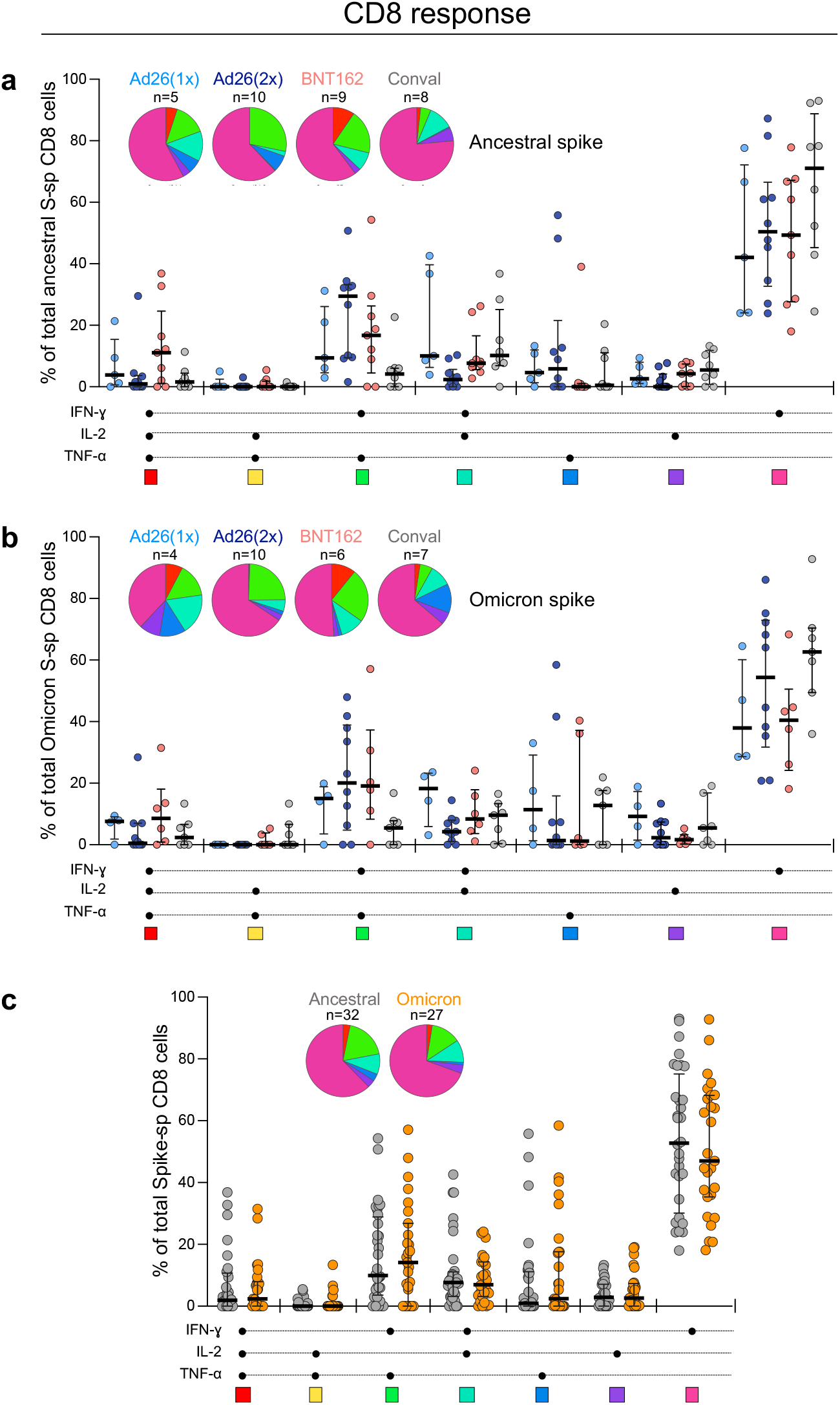
Polyfunctional profiles of SARS-CoV-2-specific CD8 T cells after vaccination and in unvaccinated convalescent volunteers. **a, b**, Comparison of the polyfunctional profile of ancestral (**a**) and Omicron (**b**) spike-specific CD8 T cells between the four groups (Ad26.COV2.S-one dose, Ad26.COV2.S-two doses, BNT162b2-two doses and unvaccinated convalescent COVID-19 volunteers). **c**, Comparison of the polyfunctional profile between ancestral spike and Omicron spike-specific CD8 T cells including all CD8 T cell responding participants, irrespective of their clinical grouping. The medians and IQR are shown. Each response pattern (i.e., any possible combination of IFN-γ, IL-2 or TNF-α expression) is color-coded, and data are summarized in the pie charts. No significant differences were observed between pies using a permutation test. The number of participants included in each analysis is indicated on the graphs.

**Extended Data Fig. 4:**
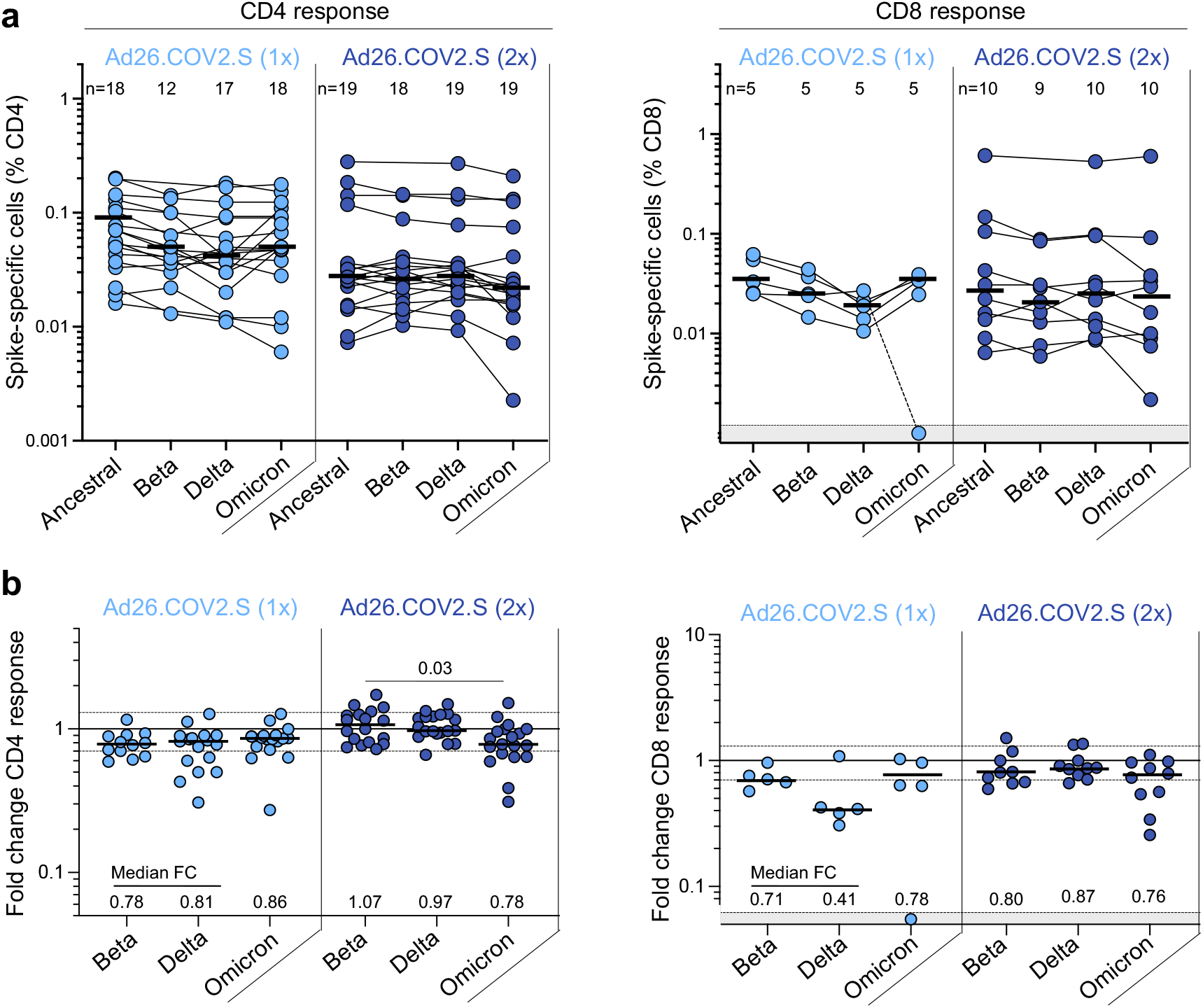
T cell responses to the ancestral, Beta, Delta and Omicron SARS-CoV-2 spike in participants who received Ad26.COV2.S (one or two doses). **a**, Frequency of spike-specific CD4 (left panel) and CD8 T cells (right panel) producing any of the measured cytokines (IFN-γ, IL-2 or TNF-α) in response to ancestral, Beta, Delta and Omicron spike peptide pools. Bars represent median of responders. No significant differences were observed between variants using a Kruskal-Wallis test with Dunn’s multiple comparisons post-test. **b**, Fold change in the frequency of spike-specific CD4 (left panel) and CD8 T cells (right panel) between ancestral and Omicron spike responses. Bars represent medians. Differences between SARS-CoV-2 variants were calculated using a Kruskal-Wallis test with Dunn’s multiple comparisons post-test. Median fold changes are indicated at the bottom of each graph. The number of participants included in each analysis is indicated on the graphs.

**Extended Data Fig. 5:**
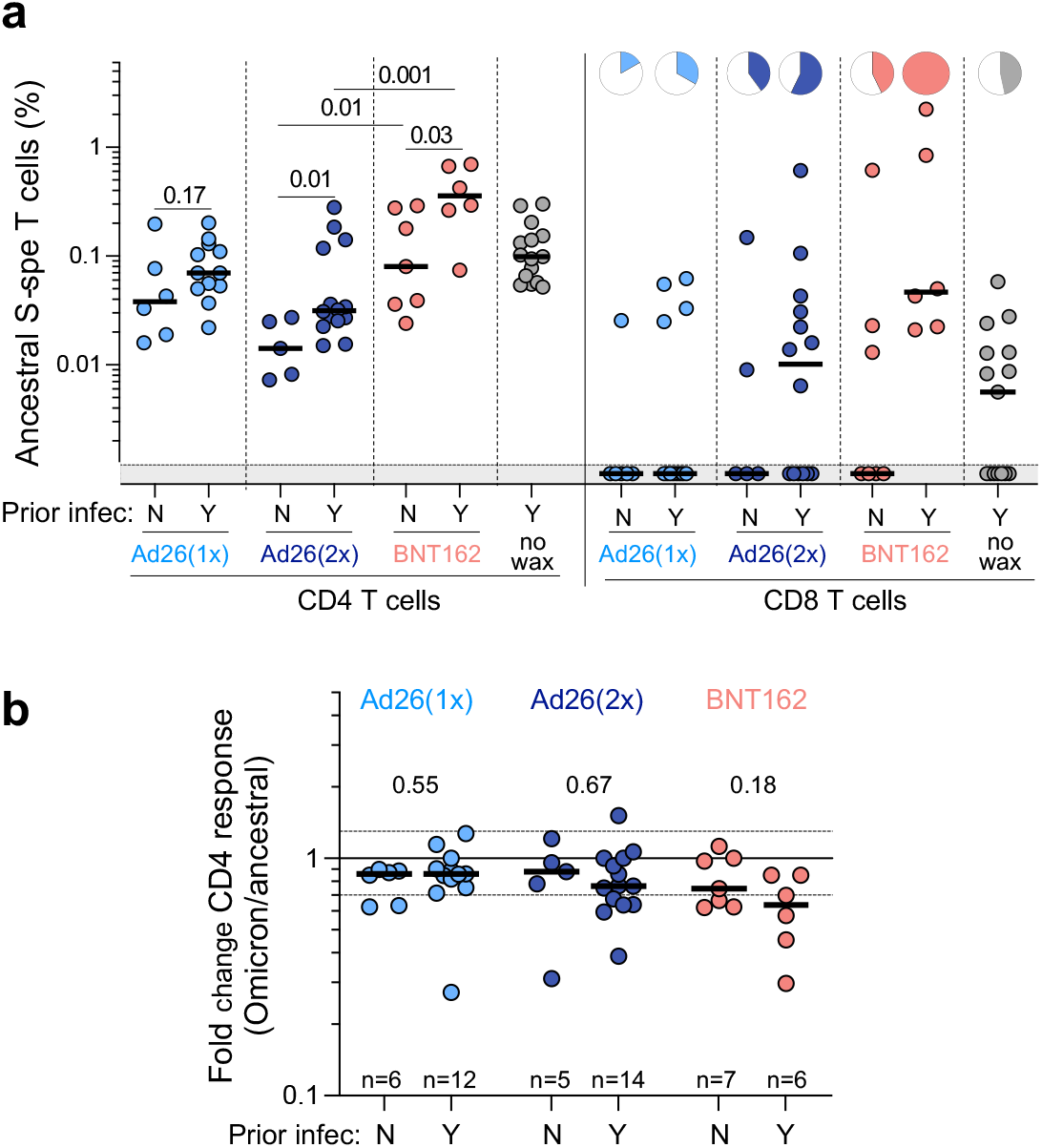
Impact of prior COVID-19 infection on T cell responses to the ancestral and Omicron SARS-CoV-2 spike in vaccinated participants. **a**, Comparison of the frequency of ancestral spike-specific T cell responses in vaccinated participants who had (Y) or did not have (N) prior SARS-CoV-2 infection. Pies depict the proportion of participants exhibiting a detectable CD8 T cell response. Bars represent medians. Statistical differences were calculated using a Mann-Whitney test. **b**, Fold change in the frequency of spike-specific CD4 T cells between ancestral and Omicron spike responses in the three vaccine groups. Bars represent medians. Statistical differences were calculated using a Mann-Whitney test. The number of participants included in each analysis is indicated on the graphs.

## METHODS

### Human Participants

At total of 138 participants were included in this study and grouped according to their vaccination and COVID-19 status. Participants were selected based on PBMC availability. The study was approved by the University of Cape Town Human Research Ethics Committee (ref: HREC 190/2020, 207/2020 and 209/2020) and the University of the Witwatersrand Human Research Ethics Committee (Medical) (ref. M210429 and M210752), the Biomedical Research Ethics Committee at the University of KwaZulu– Natal (ref. BREC/00001275/2020) and the University of Pretoria Health Sciences Research Ethics Committee (ref. 247/2020). Written informed consent was obtained from all participants.

#### 1 Participants vaccinated with Ad26.COV2.S (one or two doses) or BNT162b2 (two doses)

PBMC samples from 40 participants (20 who received one dose of Ad26.COV2.S vaccine and 20 who received two doses) were included in this study. These participants are enrolled in the Sisonke Phase 3b trial, an implementation trial of Ad26.COV2.S in healthcare workers. Recruitment took place at Groote Schuur Hospital (Cape Town, Western Cape, South Africa) between July 2020 and December 2021. Prior COVID-19 infection was recorded in 13 out of the 20 participants who had received one dose of the Ad26.COV2.S vaccine and in 14 out of 20 participants who had received two doses. Additionally, we also included samples from 15 participants vaccinated with two doses of BNT162b2 (Pfizer), enrolled in a prospective cohort study in KwaZulu Natal (South Africa). Prior COVID-19 infection was recorded for 6 out of 15 participants. The demographic and clinical characteristics of vaccinated participants are summarized in **Supplementary Table 1**.

#### 2 Convalescent COVID-19 participants

COVID-19 convalescent volunteers (n = 15) were recruited from Groote Schuur Hospital in Cape Town (Western Cape, South Africa). Based on the reported date of infection, seven were likely infected with ancestral SARS-CoV-2 (prior to August 2020), while for other 8, the infection date occurred in December 2020, suggesting an infection with the Beta variant. Samples were obtained between January 19^th^ and February 15^th^, 2021 prior to SARS-CoV-2 vaccination becoming available in South Africa. All had a documented positive SARS-CoV-2 PCR swab result or a positive SARS-CoV-2 nucleocapsid-specific antibody result (Roche Elecsys assay, Roche Diagnostics, Basel, Switzerland). The median time post positive test was 1.4 months, ranging from 1 to 7 months. The demographic and clinical characteristics of convalescent volunteers are summarized in **Supplementary Table 2**.

#### 3 Hospitalized COVID-19 patients

Sixty-eight hospitalized COVID-19 patients were included in this study. These participants were grouped according to the time of their hospitalization, reflecting four distinct infection waves in South Africa, each dominated by a different SARS-CoV-2 strain (**Fig. 2b**). Wave 1, 2 and 3 participants were recruited from Groote Schuur Hospital in Cape Town (Western Cape, South Africa) and wave 4 patients were recruited from Groote Schuur Hospital and Tshwane District Hospital in Tshwane (Gauteng, South Africa). Wave 1 patients (n = 17) were enrolled between June 11^th^ and July 24^th^, 2020, at a time when ancestral (Wuhan-1 D614G)-related SARS-CoV-2 strains were circulating. No viral sequences are available for these patients, but we assumed that all were infected with a virus closely related to the ancestral virus, as sampling occured almost three months before the emergence of the Beta variant in South Africa. Wave 2 patients (n = 16) were recruited between December 31^st^, 2020 and January 15^th^, 2021, when the Beta variant dominated. Viral sequences were available for six second wave participants, all of which were confirmed Beta infection (GISAID accession numbers: EPI_ISL_1040693, 1040658, 1040661, 1040685, 1040657, 1040663). Wave 3 patients (n = 16) were recruited between July 14^th^ and 21^st^, 2021. Wave 3 was dominated by the Delta variant. Viral sequences were available for 7 third wave participants, all of which were confirmed to be Delta infection (GISAID accession numbers: EPI_ISL_3506484, 3506367, 3957813, 3506504, 3506512, 3506518). Wave 4 patients (n = 19) were recruited between December 1^st^ and 15^th^, 2021. The SARS-CoV-2 Omicron variant was dominant during this current wave. Amongst those patients, seven had a Taqpath PCR test performed (Thermofisher, Waltham, Massachusetts, USA), all of which were characterized by a S gene target failure, highly suggestive of an Omicron infection. Moreover, swab samples were available for another five patients included in this group and whole genome sequencing is pending. The demographic and clinical characteristics of hospitalized COVID-19 participants are summarized in **Supplementary Table 3**.

### SARS-CoV-2 spike WGS and phylogenetic analysis

Whole genome sequencing (WGS) of SARS-CoV-2 was performed from nasopharyngeal swabs. Sequencing was performed as previously published^2^. Briefly, RNA was extracted on an automated Chemagic 360 instrument, using the CMG-1049 kit (Perkin Elmer, Hamburg, Germany). Libraries for whole genome sequencing were prepared using either the Oxford Nanopore Midnight protocol with Rapid Barcoding or the Illumina COVIDseq Assay. The quality control checks on raw sequence data and the genome assembly were performed using Genome Detective 1.132 (https://www.genomedetective.com) which was updated for the accurate assembly and variant calling of tiled primer amplicon Illumina or Oxford Nanopore reads, and the Coronavirus Typing Tool. Raw reads from the Illumina COVIDSeq protocol were assembled using the Exatype NGS SARS-CoV-2 pipeline v1.6.1, (https://sars-cov-2.exatype.com/). Phylogenetic classification of the genomes was done using the widespread dynamic lineage classification method from the ‘Phylogenetic Assignment of Named Global Outbreak Lineages’ (PANGOLIN) software suite (https://github.com/hCoV-2019/pangolin).

### Isolation of PBMC

Blood was collected in heparin tubes and processed within 4 hours of collection. Peripheral blood mononuclear cells (PBMC) were isolated by density gradient sedimentation using Ficoll-Paque (Amersham Biosciences, Little Chalfont, UK) as per the manufacturer’s instructions and cryopreserved in freezing media consisting of heat-inactivated fetal bovine serum (FBS, Thermofisher Scientific) containing 10% DMSO and stored in liquid nitrogen until use.

### SARS-CoV-2 antigens

For T cell assays on hospitalized patients, we used commercially available peptide pools (15mer sequences with a 11 amino acids overlap) covering the full length of the Wuhan-1 SARS-CoV-2 nucleocapsid, membrane and spike proteins (PepTivator®, Miltenyi Biotech, Bergisch Gladbach, Germany). For spike, we combined two peptide pools covering the N-terminal S1 domain of SARS-CoV-2 from aa 1 to 692 and the majority of the C-terminal S2 domain. Pools were resuspended in distilled water at a concentration of 50 µg/mL and used at a final concentration of 1 µg/mL. To determine T cell responses to SARS-CoV-2 variants in vaccinated and convalescent volunteers, we used custom mega pools of peptides. These peptides (15-mers overlapping by 10 amino acids) spanned the entire spike protein corresponding to the ancestral Wuhan sequence (GenBank: MN908947), Beta (B.1.351; GISAID: EPI_ISL_736932), Delta SARS-CoV-2 variants (B.1.617.2; GISAID: EPI_ISL_2020950) or Omicron (B.1.1.529), carrying in the spike sequence all the 38 currently described mutations (A67V, H69del, V70del, T95l, G142D, V143del, Y144del, Y145del, S152W, N211del, L212l, ins214EPE, G339D, S371L, S373P, S375F, K417N, N440K, G446S, S477N, T478K, E484A, Q493R, G496S, Q498R, N501Y, Y505H, T547K, D614G, H655Y, N679K, P681H, N764K, D796Y, N856K, Q954H, N969K, L981F). Briefly, peptides were synthesized as crude material (TC Peptide Lab, San Diego, CA). All peptides were individually resuspended in dimethyl sulfoxide (DMSO) at a concentration of 10-20 mg/mL. Megapools for each antigen were created by pooling aliquots of these individual peptides in the respective SARS-CoV-2 spike sequences, followed by sequential lyophilization steps, and resuspension in DMSO at 1 mg/mL. Pools were used at a final concentration of 1 µg/mL with an equimolar DMSO concentration in the non-stimulated control.

### Cell stimulation and flow cytometry staining

Cryopreserved PBMC were thawed, washed and rested in RPMI 1640 containing 10% heat-inactivated FCS for 4 hours prior to stimulation. PBMC were seeded in a 96-well V-bottom plate at ∼2 × 10^6^ PBMC per well and stimulated with either the commercial ancestral SARS-CoV-2 spike (S), Nucleocapsid (N) or membrane protein (M) peptide pools (1 µg/mL) obtained from Miltenyi or custom spike mega pools corresponding to the ancestral (Wuhan-1), Beta, Delta or Omicron variants (1 µg/mL). All stimulations were performed in the presence of Brefeldin A (10 µg/mL, Sigma-Aldrich, St Louis, MO, USA) and co-stimulatory antibodies against CD28 (clone 28.2) and CD49d (clone L25) (1 µg/mL each; BD Biosciences, San Jose, CA, USA). As a negative control, PBMC were incubated with co-stimulatory antibodies, Brefeldin A and an equimolar amount of DMSO. After 16 hours of stimulation, cells were washed, stained with LIVE/DEAD™ Fixable VIVID Stain (Invitrogen, Carlsbad, CA, USA) and subsequently surface stained with the following antibodies: CD14 Pac Blue (TuK4, Invitrogen Thermofisher Scientific), CD19 Pac Blue (SJ25-C1, Invitrogen Thermofisher Scientific), CD4 PERCP-Cy5.5 (L200, BD Biosciences, San Jose, CA, USA), CD8 BV510 (RPA-8, Biolegend, San Diego, CA, USA). Cells were then fixed and permeabilized using a Cytofix/Cyto perm buffer (BD Biosciences) and stained with CD3 BV650 (OKT3) IFN-γ Alexa 700 (B27), TNF-α BV786 (Mab11) and IL-2 APC (MQ1-17H12) from Biolegend. Finally, cells were washed and fixed in CellFIX (BD Biosciences). Samples were acquired on a BD Fortessa flow cytometer and analyzed using FlowJo (v10.8, FlowJo LLC, Ashland, OR, USA). A gating strategy is provided in **Extended Data Fig. 1**. Results are expressed as the frequency of CD4 or CD8 T cells expressing IFN-γ, TNF-α or IL-2. Due to high TNF-α backgrounds, cells producing TNF-α alone were excluded from the analysis. All data are presented after background subtraction.

### Live virus neutralization assay

A live neutralization assay was performed on plasma obtained from 10 out of the 15 participants vaccinated with BNT162b2 included in this study. H1299-E3 cells were plated in a 96-well plate (Corning) at 30,000 cells per well 1 day pre-infection. Plasma was separated from EDTA-anticoagulated blood by centrifugation at 500 rcf for 10 min and stored at -80 °C. Aliquots of plasma samples were heat-inactivated at 56 °C for 30 min and clarified by centrifugation at 10,000 rcf for 5 min. Virus stocks were used at approximately 50-100 focus-forming units per microwell and added to diluted plasma. Antibody-virus mixtures were incubated for 1 h at 37 °C, 5% CO2. Cells were infected with 100 μL of the virus–antibody mixtures for 1 h, then 100 μL of a 1X RPMI 1640 (Sigma-Aldrich, R6504), 1.5% carboxymethylcellulose (Sigma-Aldrich, C4888) overlay was added without removing the inoculum. Cells were fixed 18 h post-infection using 4% PFA (Sigma-Aldrich) for 20 min. Foci were stained with a rabbit anti-spike monoclonal antibody (BS-R2B12, GenScript A02058) at 0.5 μg/mL in a permeabilization buffer containing 0.1% saponin (Sigma-Aldrich), 0.1% BSA (Sigma-Aldrich) and 0.05% Tween-20 (Sigma-Aldrich) in PBS. Plates were incubated with primary antibody overnight at 4 °C, then washed with wash buffer containing 0.05% Tween-20 in PBS. Secondary goat anti-rabbit horseradish peroxidase (Abcam ab205718) antibody was added at 1 μg/mL and incubated for 2 h at room temperature with shaking. TrueBlue peroxidase substrate (SeraCare 5510-0030) was then added at 50 μL per well and incubated for 20 min at room temperature. Plates were imaged in an ELISPOT instrument with built-in image analysis (C.T.L).

### SARS-CoV-2 pseudovirus-based neutralization assay

A pseudovirus-based neutralization assay was performed on plasma obtained from all participants vaccinated with two doses of Ad26.COV2.S (n = 20). SARS-CoV-2 pseudotyped lentiviruses were prepared by co-transfecting the HEK 293T cell line with the SARS-CoV-2 614G spike (D614G) or SARS-CoV-2 Beta spike (L18F, D80A, D215G, K417N, E484K, N501Y, A701V, 242-244 del) plasmids with a firefly luciferase encoding lentivirus backbone plasmid. The parental plasmids were provided by Drs Elise Landais and Devin Sok (IAVI). For the neutralization assays, heat-inactivated plasma samples were incubated with SARS-CoV-2 pseudotyped virus for 1 hour at 37°C, 5% CO_2_. Subsequently, 1×10^4^ HEK293T cells engineered to overexpress ACE-2, provided by Dr Michael Farzan (Scripps Research Institute), were added and the incubated at 37°C, 5% CO_2_ for 72 hours, upon which the luminescence of the luciferase gene was measured. CB6 and CA1 monoclonal antibodies were used as controls.

### Statistical analysis

Statistical analyses were performed in Prism (v9; GraphPad Software Inc, San Diego, CA, USA). Non-parametric tests were used for all comparisons. The Mann-Whitney, Friedman and Wilcoxon tests were used for unmatched and paired samples, respectively. *P* values less than 0.05 were considered statistically significant. Details of analysis performed for each experiment are described in the figure legends.

## Acknowledgements

We are indebted to the participants who volunteered samples to this study. We thank Wesley van Hougenhouck-Tulleken for assistance with database management for the Pretoria COVID-19 study; Rajiev Ramlall and Jane Semugga for assistance with participant recruitment at Tshwane District Hospital. We thank Cathrine Scheepers and Josie Everatt for generous assistance with plotting lineage prevalence, and the Network for Genomic Surveillance-South Africa (NGS-SA) for providing sequencing data rapidly and in an open manner. We acknowledge the National Health Laboratory Service Greenpoint Laboratory for PCR testing. We thank Anupa Singh and Leonard Lazarus from Biocair for assistance with shipping delays of critical reagents. We acknowledge the members of the South African Variant Consortium led by Willem Hanekom and Tulio de Oliveira for discussion.

## Author contributions

W.A.B, C.R. and R.K. designed the study. C.R. and W.A.B analyzed the data and wrote the manuscript. K.K., S.C., M.B., F.K. and S.V.M. performed neutralization assays and analyzed the data. R.K., M.B.T., A.N., R.B., N.B., and A.S. performed flow cytometry assays. M.M., S.S., M.A., D.M., O.A., C.S., E.dB., M.A.vdM., Z.dB., T.R.dV. A.B., G.vdB, A.M., A.S., M.V. recruited patients, managed the cohorts and contributed clinical samples and data. A.G., D.W. and A.S. designed and provided variant peptide pools. T.M-G., J.B, P.L.M. and A.S designed, supervised and analysed the neutralization assay. N.A.B.N., R.J.W., V.U., M.T.B and T.R. established and led the clinical cohorts and contributed samples. L.G.B and G.G. led the Sisonke Ad26.COV2.S vaccination trial. All authors reviewed and edited the manuscript.

