## Supplemental Tables for "SARS-CoV-2 spike T cell responses induced upon vaccination or infection remain robust against Omicron"

**Supplementary Table 1**

| Vaccinee cohorts | Vaccinees (all) | Without prior infection | With prior infection |
|---|---|---|---|
| **Ad26.COV2.S (Janssen) - 1 dose** | | | |
| Number | 20 | 7 | 13 |
| Age (y) (median, IQR) | 41 [32-52] | 44 [32-54] | 41 [32-48] |
| Gender (% female) | 75% | 71.4% | 76.9% |
| With co-morbidities (n, %) | 6/20 (30%) | 2/7 (28.6%) | 4/13 (30.8%) |
| Days since last vaccine dose (median, IQR) | 28 [28-32] | 31 [28-34] | 28 [27-29] |
| Months since COVID episode (median, IQR) | - | na | 3 [2-6]* |
| **Ad26.COV2.S (Janssen) - 2 doses** | | | |
| Number | 20 | 6 | 14 |
| Age (y) (median, IQR) | 38 [33-56] | 58 [36-62] | 36 [32-44] |
| Gender (% female) | 75% | 83.3% | 71.4% |
| With co-morbidities (n, %) | 9/20 (45%) | 4/6 (66.7%) | 5/14 (35.7%) |
| Days since last vaccine dose (median, IQR) | 22 [21-24] | 23 [21-25] | 22 [21-23] |
| Months since COVID episode (median, IQR) | - | na | 15 [11-18] |
| **BNT162b2 (Pfizer) - 2 doses** | | | |
| Number | 15 | 9 | 6 |
| Age (y) (median, IQR) | 53 [45-71] | 53 [41-74] | 57 [44-66] |
| Gender (% female) | 53.3% | 44.4% | 66.7% |
| With co-morbidities (n, %) | 9/15 (60%) | 4/9 (44.4%) | 5/6 (83.3%) |
| Days since last vaccine dose (median, IQR) | 33 [22-48] | 46 [23-65] | 27 [20-38] |
| Months since COVID episode (median, IQR) | - | na | 14 [13-15] |

**Supplementary Table 1: Clinical characteristics of vaccinee cohorts.**
Co-morbidities include: asthma, hypertension, obesity or diabetes mellitus.
*: data regarding time post Covid-19 infection were available for only 6 out of the 13 participants who received 1 dose of Ad26-COV.S.

**Supplementary Table 2**

| Convalescent cohort (unvaccinated) | |
|---|---|
| Number | 15 |
| Age (y) (median, IQR) | 39 [35-49] |
| Gender (% female) | 12% |
| With co-morbidities (n, %) | 6/15 (40%) |
| Months since COVID episode (median, IQR) | 1.4 [1.3-6] |

**Supplementary Table 2: Clinical characteristics of convalescent COVID-19 patients.** Co-morbidities include: asthma, hypertension, obesity or diabetes mellitus.

| Hospitalized COVID-19 cohort | Total | Wave 1 (Ancestral) | Wave 2 (Beta) | Wave 3 (Delta) | Wave 4 (Omicron) |
|---|---|---|---|---|---|
| Number | 68 | 17 | 16 | 16 | 19 |
| Age (y) (median, range) | 53 [39-64] | 55 [41-62] | 64 [53-68] | 49 [37-63] | 47 [35-60] |
| Gender (% female) | 47% | 29.4% | 56.2% | 56.2% | 47.4% |
| With co-morbidities (n, %) | 53/68 (77.9%) | 11/17 (64.7%) | 15/16 (93.7%) | 13/16 (81.2%) | 14/19 (73.7%) |
| Day since PCR+ (median, range) | 4 [2-8] | 2 [1-3.5] | 5 [2-10] | 6 [2-10] | 4 [3-6] |
| Severe COVID-19 (WHO ≥5) | 26/68 (38.2%) | 7/17 (41.2%) | 1/16 (6.25%) | 9/16 (56.2%0 | 9/19 (47.4%) |

**Supplementary Table 3: Clinical characteristics of hospitalized COVID-19 patient cohort.**
Co-morbidities include: asthma, hypertension, obesity or diabetes mellitus. Severe COVID-19 was defined based on oxygen therapy requirement according to the WHO ordinal scale scoring system ($O_2$ via high flow to extracorporeal membrane oxygenation).
